# IL-13 associated epithelial remodeling correlates with clinical severity in nasal polyposis

**DOI:** 10.1101/2022.10.06.22280802

**Authors:** Maya E. Kotas, Neil N. Patel, Emily K. Cope, Jose G. Gurrola, Andrew N. Goldberg, Steven D. Pletcher, Max A. Seibold, Camille M. Moore, Erin D. Gordon

**Author notes:** co-corresponding authors Corresponding authors: Erin D. Gordon, MD, Division of Pulmonary, Critical Care, Allergy and Sleep Medicine, Department of Medicine, University of California, San Francisco, 513 Parnassus Ave, HSE 201, San Francisco, CA 94143, Camille M. Moore, PhD, Center for Genes, Environment, and Health, National Jewish Health, 1400 Jackson St., Denver, Colorado 80206. **Declaration of interests** JGG is a Genentech Advisory Board Consultant with agreement ending December 2021. ANG is a minor stock holder in Siesta Medical. SDP and ANG are co-inventors of patent 14/394, 006 Sinus diagnostics and treatments. The other authors declare no competing interests.

## Abstract

**Background:** Epithelial remodeling is a histopathologic feature of chronic inflammatory airway diseases including chronic rhinosinusitis (CRS). Cell type shifts and their relationship to CRS endotypes and severity are incompletely described.

**Objective:** The purpose of this study was to understand the relationship of epithelial cell remodeling to inflammatory endotypes and disease outcomes in CRS.

**Methods:** Using cell type transcriptional signatures derived from epithelial single cell sequencing, we analyzed bulk RNA sequencing data from sinus epithelial brushes obtained from patients with CRS with and without nasal polyps in comparison to healthy controls.

**Results:** The airway epithelium in nasal polyposis displayed increased tuft cell transcripts and decreased ciliated cell transcripts along with an IL-13 activation signature. In contrast, chronic rhinosinusitis without polyps showed an IL-17 activation signature. IL-13 activation scores were associated with increased tuft cell, goblet cell and mast cell scores and decreased ciliated cell scores. Furthermore, the IL-13 score was strongly associated with a previously reported activated (“polyp”) tuft cell score and a prostaglandin E2 (PGE2) activation signature. The Lund-McKay score, a computed tomographic metric of sinus opacification, correlated positively with activated tuft cell, mast cell, PGE2, and IL-13 and negatively with ciliated cell transcriptional signatures.

**Conclusions:** These results demonstrate that cell type alterations and PGE2 stimulation are key components of IL-13 induced epithelial remodeling in nasal polyposis, while IL-17 signaling is more prominent in CRS without polyps, and that clinical severity correlates with the degree of IL-13 induced epithelial remodeling.

**Key Messages:**

- Cell type signatures from single cell RNA sequencing, applied to bulk sequenced RNA sinus brushes, suggest increased tuft cells and mast cells and decreased ciliated cells in nasal polyp epithelium.
- IL-17 signaling rather than IL-13 signaling is observed in epithelium from CRSsNP.
- IL-13-drives epithelial remodeling and prostaglandin E2 signatures correlated with clinical measures of sinus opacification in CRS.

**Capsule Summary:** Measures of epithelial remodeling, including both IL-13 and PGE2 induced epithelial activation and cell type specific transcript alterations, correlate with a radiographic metric of disease severity in CRSwNP.

## Introduction

Chronic rhinosinusitis (CRS) is a chronic inflammatory condition affecting the nasal mucosa and paranasal sinuses in 10% of the adult population (1). Although patients suffer a common array of symptoms (including congestion, nasal discharge, facial pressure and anosmia), CRS is increasingly recognized to be a heterogenous disease. Two major forms of CRS are recognized: CRS with nasal polyps (CRSwNP; referred herein as “polyp” for simplicity) or without polyps (CRSsNP). Within these major subtypes, further pathologic or endotypic divisions are incompletely defined. Epithelial remodeling, epithelial barrier dysfunction, shifts in the microbial ecology, and immune activation are key processes that are thought to contribute to the pathophysiology, but the specific sequence of these events or relationship to one another are not known.

Critical to developing or selecting therapies for patients with chronic inflammatory airway diseases like CRS are characterizing the epithelial changes (“remodeling”) that drive clinical pathology, and understanding how this remodeling is directed by inflammatory mediators. IL-13 is widely accepted to be a key driver of goblet cell differentiation in type 2 inflammation, but other epithelial changes and inflammatory mediators are less well understood. Additionally, existing data on cell type composition changes that might be driven by inflammation are inconsistent. For instance, basal cells have been reported in some publications to be increased in polyps (2), but this finding has not been consistent (3). Moreover, the airway epithelial landscape includes rare and less-understood populations of epithelial cells such as ionocytes, neuroendocrine cells, and tuft cells, which likely modify mucociliary and immune behaviors but have not been well characterized in the setting of airway inflammation.

Traditionally, studies of epithelial remodeling in chronic airway diseases such as CRS have relied on typical morphology or immunofluorescence staining to identify and enumerate cell types of interest. More recently, single cell RNA sequencing (scSeq) has identified previously-undiscovered populations and enabled more precise definitions of known cell types based on concurrent expression of multiple unique or enriched transcriptional markers. While scSeq has improved airway cell type definitions, application of the technique to clinical specimens remains challenging due to high costs, differential ability of distinct cell types to survive tissue digestion protocols (leading to under-representation), and tendency to miss rare cell types when sequencing small-to-moderate numbers of cells.

In a prior publication, we used scSeq to compare epithelial brushings from 5 nasal polyps to 4 healthy sinuses. We discovered that polyp brushings had increased numbers of tuft cells newly expressing eicosanoid synthetic enzymes, and an associated PGE2-induced transcriptional signature across the polyp epithelium. By applying these novel transcriptional signatures to a larger cohort of banked bulk RNA from sinus epithelial brushings from 24 patients with CRSwNP, 7 patients with CRSsNP and 8 healthy control subjects, we determined that these scSeq-derived transcriptional signatures could be effectively applied to bulk RNA samples to confirm and extend our findings in a distinct patient cohort (3). Here we examine additional cell type and stimulation-state transcriptional signatures in those specimens in order to more broadly examine epithelial remodeling in CRS. We find that transcriptional signatures of IL-13 correlate with reductions in ciliated cells, increases in tuft cells, goblet cells and mast cells, epithelial prostaglandin E2 (PGE2) activation, and radiographic severity scores in patients with CRSwNP. By contrast, we find that CRSsNP is associated with IL-17 stimulation without alterations in cell type composition.

## Methods

### Sinus Study Participants

Subjects between the ages of 18 and 75 years (previously described in (3)) were recruited from the University of California, San Francisco Hospital (San Francisco, California) Otolaryngology clinic between 2013 and 2019 to participate in a UCSF Sinus Tissue Bank. The UCSF Committee on Human Research approved the study, and all subjects provided written informed consent. Cytologic brushes were collected from ethmoid sinus at the time of elective endoscopic sinus surgery from patients with physician-diagnosed chronic rhinosinusitis with or without nasal polyps on the basis of established guidelines. Subjects with cystic fibrosis were excluded from the study. Non-CRS control subjects were those who were undergoing endoscopic surgery for alternative indications and brushes were primarily collected from the maxillary sinuses.

### Human Biospecimen Processing

As described in (3), cytologic brushes were placed in RNAlater (Qiagen) until RNA extraction. These were defrosted on ice, diluted with sterile PBS, centrifuged, and brushes were removed and placed into lyse E tube. Pellets were resuspended in RLT/BME and added to the Lyse E tube. Samples were agitated in a bead beater for 30 sec. Samples were centrifuged at 2000 rpm for 1 min and transferred to an Allprep (Qiagen) spin column. RNA and DNA were prepared according to the manufacturer’s instructions. Residual DNA was removed from the purified RNA by incubation with RNase-Free DNase (Promega) for 30 minutes at 37°C. DNase was removed from the preparation via a second RNA clean up using the Qiagen RNeasy Kit. RNA concentration was determined using Nanodrop (Thermo Scientific) and RNA quality was assessed using Agilent Pico RNA kit.

### Bulk RNA-seq for human biospecimens

For whole transcriptome sequencing we first used the Ion AmpliSeq™ Transcriptome Human Gene Expression Kit (Cat #A26325, Life Technologies) to enable gene-level expression analysis from small amounts of RNA. We generated barcoded sequencing libraries per the manufacturer’s protocol from 10 ng of RNA isolated from the 24 stimulation samples detailed above (12 pairs).

Libraries were sequenced using the Ion PI template OT2 200 kit v3 for templating and the Ion PI sequencing 200 kit v3 kit for sequencing. Barcoding allowed all 24 samples to be loaded onto 3 PI sequencing chips and sequenced with an Ion Proton Sequencer using standard protocols. Read mapping was performed with the TMAP algorithm on the Proton server and read count tables for each gene amplicon generated using the Proton Ampliseq plugin. Read counts for gene amplicons across all 3 runs were merged to generate the final raw expression data.

### Bulk Gene Signature Scores and Comparisons

Cell-type signatures were developed based on marker lists from published scSeq studies. To generate scores, RNA-seq count data were first normalized using DESeq2’s variance stabilizing transformation (4). Normalized gene expression was then centered and scaled and a score for each sample was generated by taking the average of scaled, normalized expression for all genes in the signature.

Epithelial cell type signatures were based on cell type markers identified from scSeq of samples from polyp (N=5) and control (N=4) sinus brushings (3), which included basal cells, secretory cells, goblet secretory cells, ciliated cells, ionocytes, and tuft cells. To identify a set of cell type markers that was consistent across all subjects, we performed the following differential expression analyses. First, to identify potential cell type markers, we carried out pairwise differential expression analysis, comparing log-normalized expression in each cell type to all others using a Wilcoxon rank sum test. Potential markers were identified as genes exhibiting significant upregulation when compared to all other cell types, defined by having a Bonferroni adjusted p-value < 0.05, a log fold change > 0.25, and >10% of cells with detectable expression. This analysis was then performed separately for each subject using the FindConservedMarkers function in the Seurat v. 3 R package (5) to determine if the potential markers were consistent across subjects. Final cell-type markers were required to have significant upregulation in all nine subjects. The top 50 of these markers (ranked by maximum p-value across subjects) for each cell type were included in the gene signature.

A neuroendocrine cell signature was based on markers from Goldfarbmuren et al (6) comparing gene expression in neuroendocrine cells to other rare epithelial cell types, including ionocytes and tuft cells. Scores for mast cells and neutrophils were derived from published marker lists from Ordovas-Montanes et al (2) and Travaglini et al (7), respectively. PGE2, IL-13 and polyp tuft cell signatures were generated from the IL-13 and PGE2 pan-epithelial response genes identified in Kotas et al (3). The IL-17 score was based on Christenson et al (8).

For full lists of genes included in each score see Supplementary Table 2.

### Modeling of Gene Signature Scores

Gene signature scores were compared between patients with polyp, CRSsNP, and healthy controls using linear regression. Unadjusted models and models adjusting for patient age, sex, smoking status (ever vs. never) and race (white vs. other races) were fit. P-values from unadjusted models are shown in figures, while results from adjusted models are provided in the supplementary material. The Benjamini-Hochberg method (9) was used to correct p-values for multiple comparisons in both unadjusted and adjusted models.

Associations between the IL-13 and PGE2 response signature scores and other gene expression scores were evaluated using linear regression models. Unadjusted models and models adjusting for patient group, age, sex, smoking status and race were fit. Similar linear models were used to determine the association between Lund-Mackay (LM) scores and gene signature scores. P-values from unadjusted models are shown in figures, while results from adjusted models are provided as supplementary material. Again, the Benjamini-Hochberg method was used to correct p-values for multiple comparisons in both the unadjusted and adjusted models.

### Differential Expression and Gene Set Enrichment Analysis

Differential expression between groups was performed using DESeq2. Models controlled for age, sex, smoking status and race. P-values were corrected using the Benjamini-Hochberg method to control the false discovery rate. Gene set enrichment analysis was performed separately for up and down regulated differentially expressed genes (FDR adjusted p-value < 0.05) using the EnrichR R package (10-12) with the KEGG 2019 (13; 14) database.

### Study Approval

Sampling from human subjects was approved by the UCSF Committee on Human Research, and all subjects provided written informed consent.

## Results

To broadly examine cell type changes in CRS, we generated lists of the top 50 transcriptional markers of epithelial cell types found in healthy or polyp sinus epithelium from single cell RNA seq data (3), and averaged the scaled and normalized expression of these markers to create cell type “scores.” These included well-described and common cell types such as basal cells, goblet secretory cells, and ciliated cells, as well as rare subsets including tuft cells, neuroendocrine cells, and ionocytes. We then applied these cell type “scores” to sequenced bulk RNA-seq data obtained from our larger cohort. (Figure 1A). We observed no difference in basal cell or goblet secretory cell scores between groups (Figure 1B-C), but found that ciliated cell associated transcripts were significantly decreased in polyp patients compared to CRSsNP, and trended to decrease in polyp compared to controls (Figure 3D). Next, examining rare cell types, we detected an increase in tuft cells transcripts in polyp (Figure 1E), consistent with prior reports and our prior single cell sequencing data (3; 15). In contrast, neuroendocrine cells and ionocytes, which may share a lineage relationship with tuft cells (6), trended towards decrease in polyp patients compared to CRSsNP.

**Figure 1:**
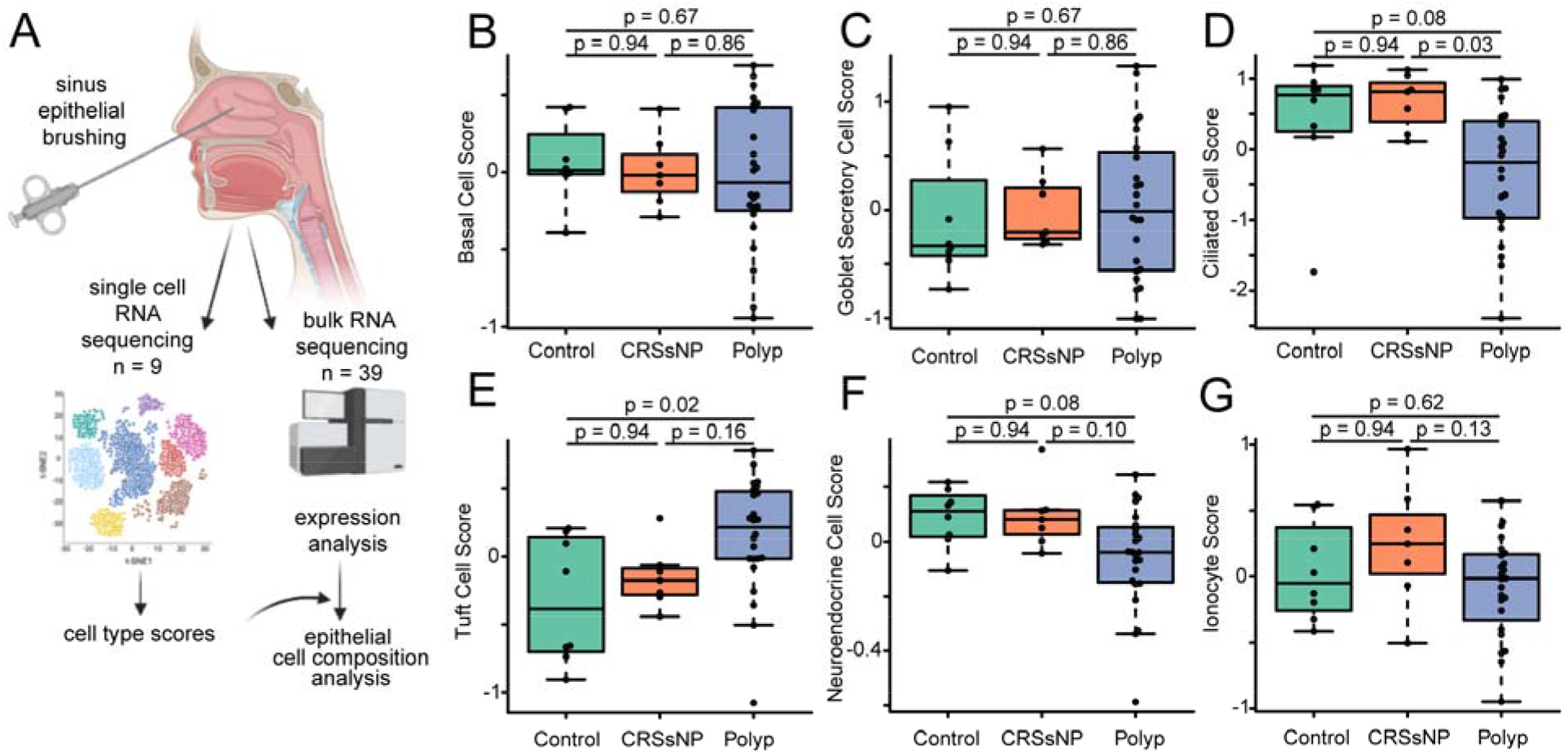
Bulk RNA sequencing suggests loss of ciliated cells and increase in tuft cells in CRSwNP. (**A**) Cell type signatures generated from single cell RNA sequencing were applied to 39 bulk RNA-sequenced endoscopic sinus brushings. (**B**) basal cell, (**C**) goblet secretory cell,(**D**) ciliated cell, (**E**) tuft cell, (**F**) neuroendocrine cell, and (**G**) ionocyte cell scores in sequenced sinus cytobrushings.

Nasal polyposis is associated with type 2 inflammation (1). We previously found that a 3-gene score described as a bronchial epithelial biomarker of the type 2-high variant of asthma (16) was increased in polyp compared to control or CRSsNP tissue (3). We reasoned that an expanded list of IL-13-responsive genes specifically identified in sinonasal tissue may better discriminate between patients with different endotypes of CRS. We analyzed expression of a set of IL-13-responsive genes that we previously found to be commonly upregulated in most epithelial cell types in polyps compared to control tissue, and found these to be significantly increased in bulk sequenced polyp epithelium, but not CRSsNP, compared to control tissue (Figure 2A-B). Consistent with this finding and the prominent involvement of mast cells in type 2 airway inflammation, we also found a significant increase in mast cell-associated transcripts in polyp but not CRSsNP epithelium compared to controls (Figure 2C).

**Figure 2:**
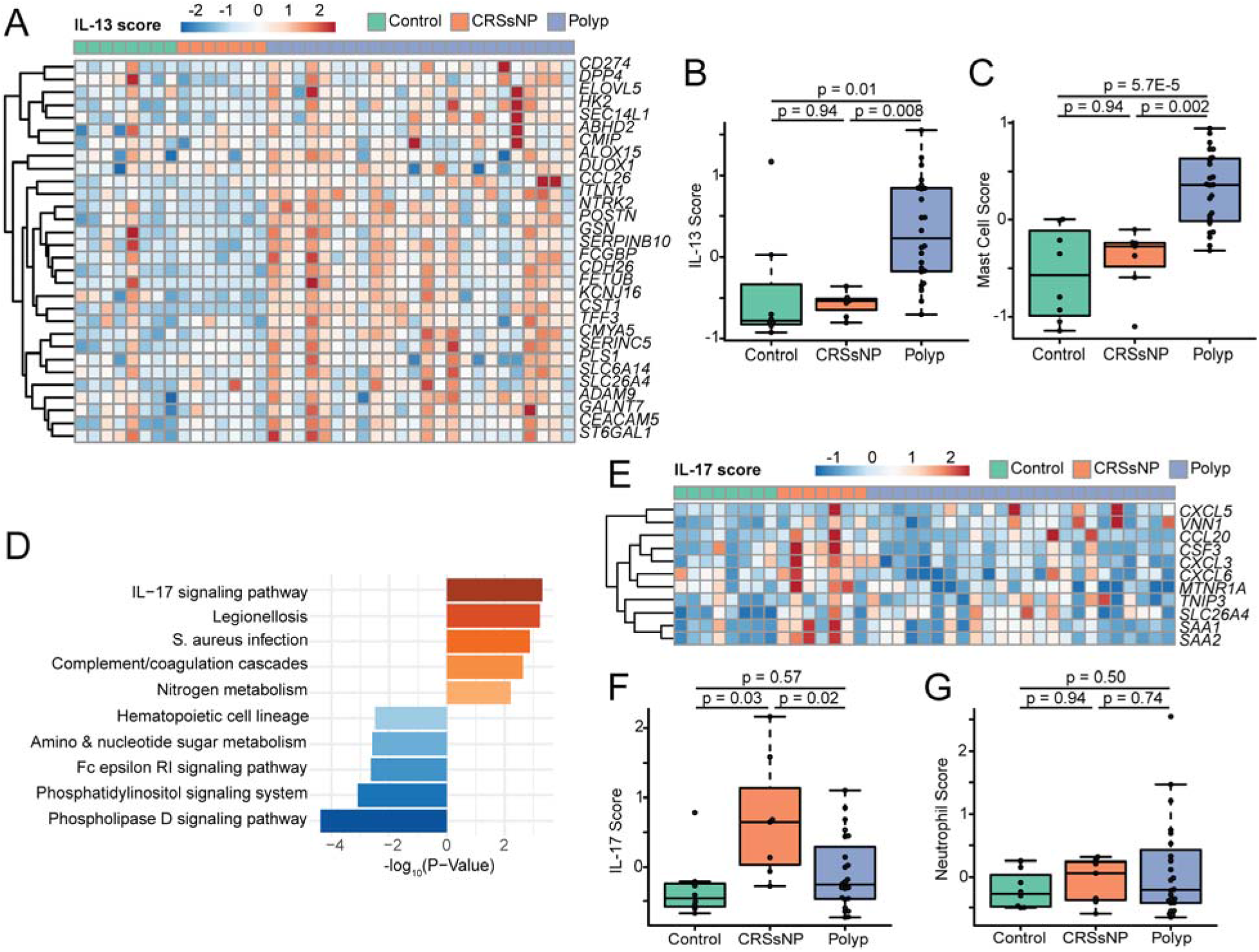
IL-13-driven inflammation dominates in nasal polyps, while IL-17-driven inflammation is found in CRSsNP. (**A**) IL-13-responsive genes are increased in polyp epithelium compared to CRSsNP or controls. (**B**) composite “score” of genes in panel (A). (**C**) Increased expression of transcripts associated with mast cells in nasal polyps. (**D**) Enrichment in IL-17 signaling in CRSsNP compared to polyp. (**E**) IL-17 response signature is elevated in CRSsNP. F. Neutrophil signatures are unaltered in CRS.

CRSsNP is known to be an inflammatory state, but our samples lacked signs of type 2 inflammation (3-gene score (16) or expanded sinus IL-13 score). Therefore, we hypothesized that a different inflammatory signature might be dominant in some patients with CRSsNP. Comparing transcripts upregulated in CRSsNP over polyps, we observed enrichment of genes associated with IL-17 signaling (Figure 2D). Because the IL-17 pathway can affect a broad array of cell types, with variable functional or transcriptional impacts, we then made use of an 11-gene IL-17-response transcriptional signature previously validated specifically in airway epithelium (8). We found that this IL-17 signature was significantly increased in epithelial brushings from patients with CRSsNP when compared to either polyp patients or controls (Figure 2E-F).

Although IL-17 signaling is sometimes associated with neutrophil recruitment, we observed no increase in neutrophil-associated transcriptional scores between patient endotypes (Figure 2G). IL-13 is thought to be a key driver of airway epithelial remodeling and cell type compositional changes (3; 17). To determine how cell type compositional changes relate to inflammatory cues in CRS, we examined the relationship between our IL-13 sinus epithelial score and the epithelial cell type scores described in Figure 1. As we expected, IL-13-associated transcripts strongly correlated with tuft cell signatures (Figure 3A). By contrast, the IL-13 score was inversely correlated with ciliated score markers. The IL-13 score also correlated strongly with makers of both goblet secretory cells and mast cells (Figure 3C-D), consistent with prior mechanistic studies (18; 19).

**Figure 3:**
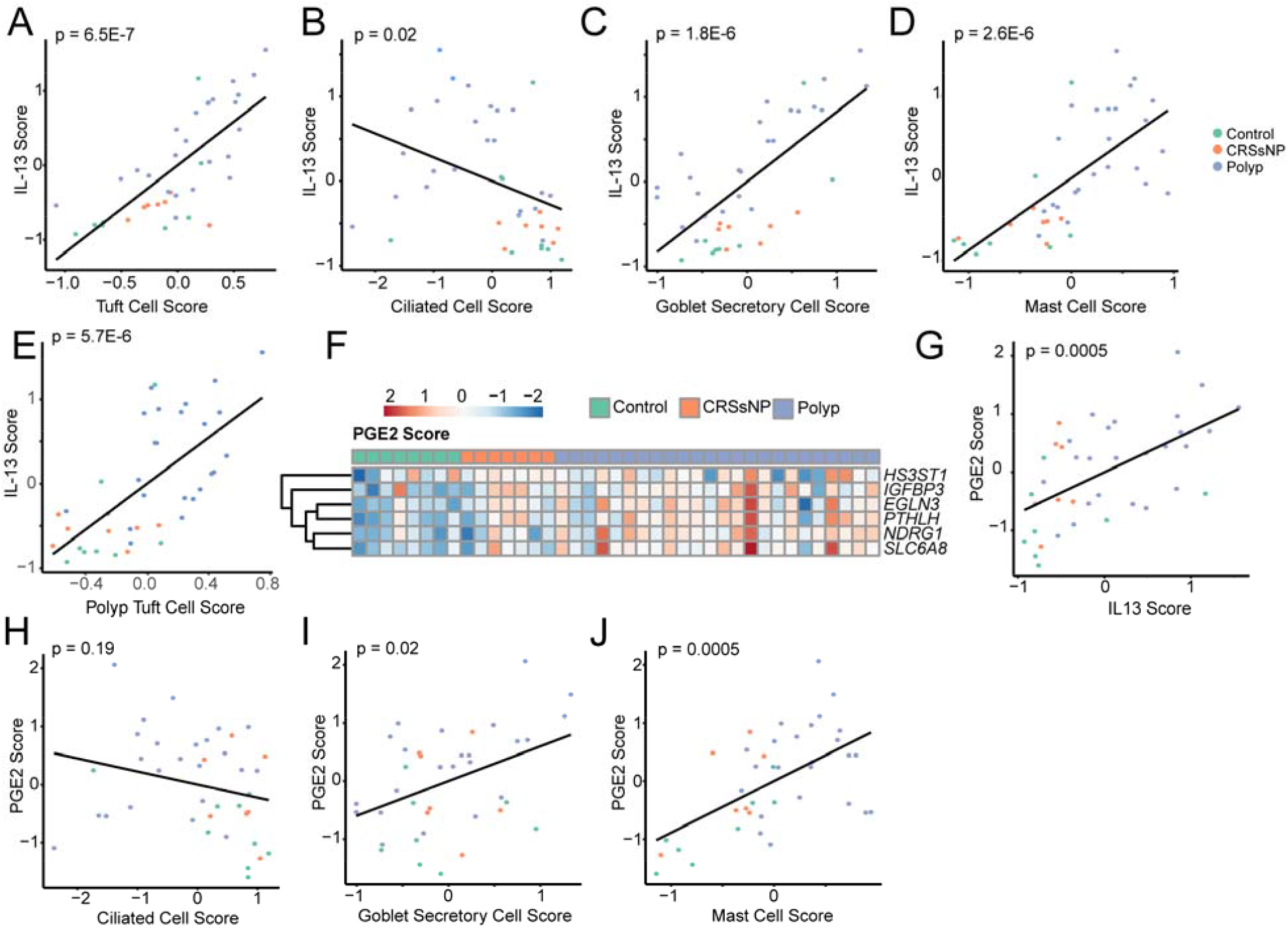
IL-13 and PGE2 signatures are associated with epithelial remodeling in CRS. Correlation of IL-13 score with transcripts associated with (**A**) tuft cells, (**B**) ciliated cells, (**C**) goblet secretory cells, and (**D**) mast cells. (**E**) Correlation of previously-described polyp tuft cell score with IL-13 score. (**F**) Increase in PGE2-response genes in CRS. (**G**) Correlation between IL-13 score and PGE2 score. Correlation between PGE2 and (**H**) ciliated, (**I**) goblet secretory and (**J**) mast cell scores in sinus.

We previously observed that tuft cells in nasal polyps adopt a novel gene expression signature (“polyp tuft cell” score), which likely enables increased production of prostaglandin E2 (PGE2), alongside a pan-epithelial transcriptional signature of PGE2 stimulation across polyp epithelium (3). Consistent with these prior findings, the PGE2 stimulation score was increased in CRS–particularly in polyps—and correlated strongly with the IL-13 score (Figure 3F-G). Because IL-13 likely drives production of PGE2 from activated tuft cells, we considered the possibility that cellular composition changes in CRS could be directed in part through PGE2. The PGE2 score poorly correlated with the ciliated cell score (Figure 3H), but was positively correlated with goblet secretory cell and mast cell transcripts (Figure 3I-J).

Clinical manifestations of CRS such as nasal congestion, mucus discharge and recurrent infections may be caused by epithelial remodeling (for instance, favoring mucus production at the expense of ciliated cells) and/or by the infiltration of immune cells which modify tissue functions through release of paracrine mediators such as cytokines. To determine how the observed cell type changes and inflammatory scores relate to clinical disease severity, we calculated Lund-Mackay (LM) scores, which measures the degree of sinus opacification based on computed tomography imaging (20). We found that the LM score was strongly positively associated with the polyp tuft cell score (Figure 4A) and strongly negatively correlated with the ciliated cell score (Figure 4B), suggesting potential contributions of disordered cell type composition to disease manifestations. Examining indicators of inflammation, we observed that the PGE2 score, IL-13 score, and mast cell scores were also correlated with LM scores (Figure 4C-D), though the IL-17 score was not.

**Figure 4:**
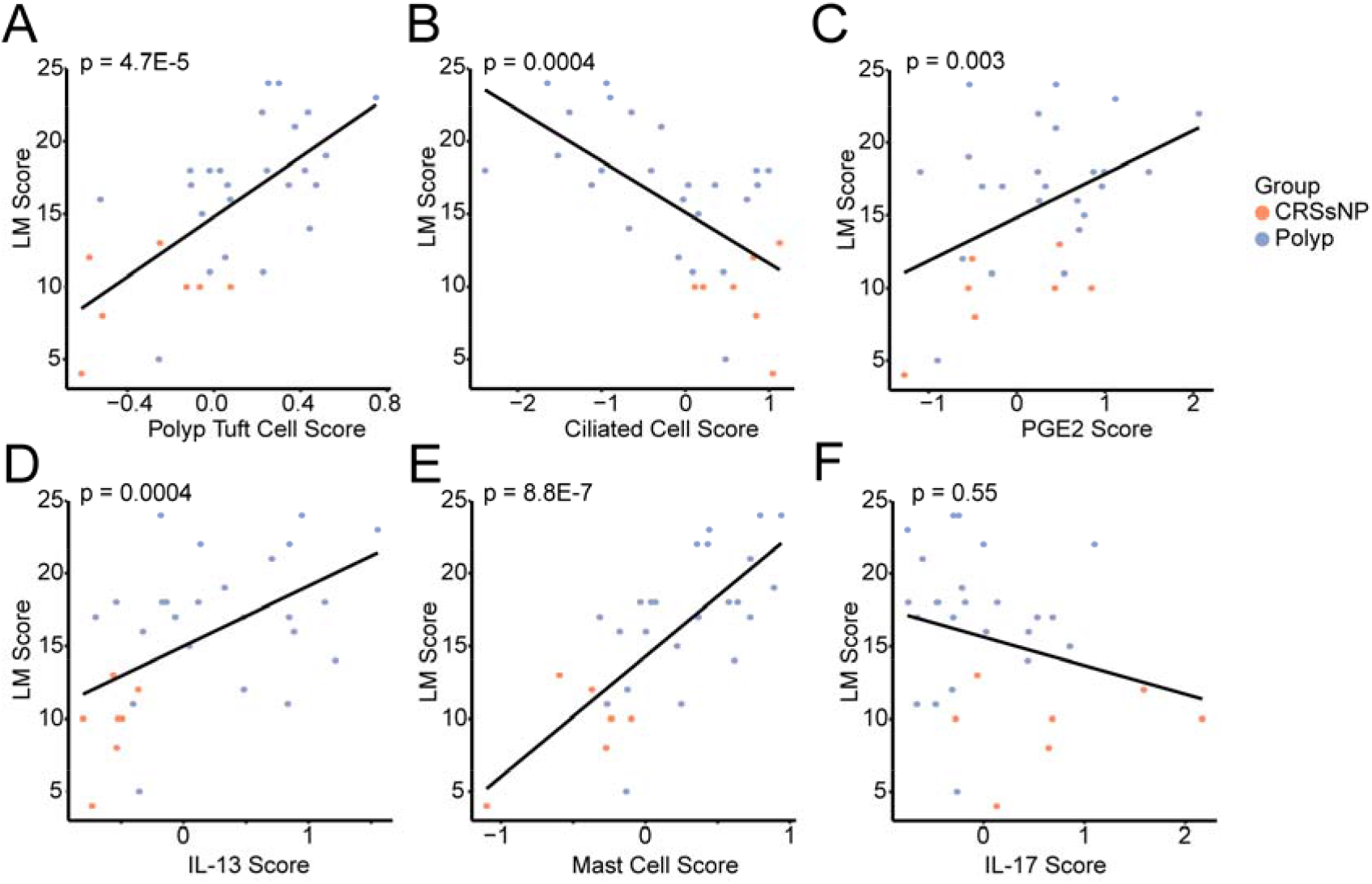
Cell type composition changes and inflammatory scores correlate with sinus opacification in CRS. Correlations between Lund-Mackay (LM) score and (**A**) polyp tuft cell score, (**B**) ciliated cell score, (**C**) PGE2 score, (**D**) IL-13 score, (**E**) mast cell score, and (**F**) IL-17 score in CRS epithelial brushes.

## Conclusions

Here we report that epithelial brushings from CRS with nasal polyps showed reduced ciliated cell-associated transcripts and increased tuft cell-related transcripts, suggesting cell type composition shifts. These cell type changes correlate with signatures of IL-13 and PGE2-driven inflammation, as well as with radiographic metrics of disease severity. By contrast, CRSsNP was associated with IL-17-driven inflammation, but this was not associated with specific cell type changes or radiographic scores. These findings suggest that clinical outcomes in CRS may be related to alterations in airway epithelial function, particularly those associated with IL-13 activity.

Our finding of decreased ciliated cell transcripts in association with IL-13 activity and worse sinus pathology is consistent with published clinical literature linking inherited or acquired ciliopathies to nasal polys and other sinus diseases (21-23) and the inhibitory effect of IL-13 on ciliary cell development and function *in vitro* (24). However, the effects of IL-13 on alterations in mucociliary function extend beyond the ciliated cells. The robust increase in goblet cells and a shift from MUC5B to MUC5AC production following IL-13 activation of the epithelium is well established (18). Consistent with this, we find a strong positive correlation between the goblet secretory score and the IL-13 activation score in the sinus epithelium. Surprisingly, the goblet secretory score was not increased in the epithelium of CRSwNP compared to health. This may be due to the small sample size or differences in anatomic location. Among control subjects, brushes were predominantly collected from the maxillary sinus, while CRS patients were all sampled from the ethmoid. Prior reports describe an increase in goblet cells within the maxillary sinus compared to ethmoid (25).

We previously showed that tuft cells increase in the epithelium of nasal polyps and that IL-13-activated tuft cells contribute significantly to airway PGE2 production (3). Others have demonstrated that tuft cell production of cysteinyl leukotrienes can augment airway type 2 inflammation (26). Our findings here suggest that tuft cell eicosanoid production may contribute to CRS pathology, as radiographic sinus opacification correlated strongly with transcriptional signatures of IL-13, tuft cell activation, and PGE2 production. While cysteinyl leukotrienes are known to augment type 2 inflammation through direct action on Th2 and ILC2 cells (27-29), PGE2 has broad physiologic effects including well described anti-inflammatory effects on Th2 and ILC2 cells (30; 31). Its tissue effects include the ability to elicit potent interstitial edema (32; 33) and activate epithelial fluid secretion via CFTR (3), which together may act to impair normal sinus drainage. PGE2 may also stimulate goblet cell development and mucin secretion, further enhancing the effects of IL-13 on goblet cell differentiation (34-36).

While we find that samples from nasal polyps have increased evidence of IL-13 stimulation, this was not evident in CRSsNP. This contrasts with prior reports suggesting that a subset of patients CRSsNP have tissue type 2 inflammation (37). In large cohort of 240 subjects with CRSsNP, Delemarre et al. measured tissue concentrations of IL-4 and IL-5 along with tissue and blood eosinophils. About half of the subjects they profiled had tissue IL-5 concentrations ≥12.98 pg/g, along with increased peripheral eosinophils, and were designated as “type 2 CRSsNP.” Only 30% of those “type-2 CRSsNP” patients had a co-morbid diagnosis of asthma. However, the lack of a control group precludes assessment of whether the measured cytokines were actually elevated when compared to healthy tissue, a critical question as we note that some of our control patients without sinus disease had evidence of type 2 inflammation. Never the less, differences between their study and ours could be explained by sample size and clinical characteristics. In our small sample of patients with CRSsNP, none had co-morbid asthma and 4/7 were current or former smokers (3).

We did find that patients with CRSsNP have evidence of IL-17 activation of the epithelium, a signature which was also present in several of the nasal polyp patients. This finding is supported by the work of Delemarre et al., which finds increased IL-17 cytokine levels in “non-type 2 CRSsNP” tissue, and suggests that IL-17 mediated inflammation is likely to be an important contributor to non-type 2 mediated CRS. Future studies are needed to determine if dysregulation of the IL-17 pathway may be mediated by bacterial products and if it might be amenable to therapeutic blockade with IL-17 directed therapeutics. While we did not see correlations between the IL-17 scores and epithelial cell type shifts or Lund-MacKay scores, our study is limited by small sample size, and such correlations are worthy of future study in larger patient cohorts.

In sum, this study provides evidence that IL-13-induced airway epithelial remodeling in nasal polyposis encompasses both the loss of ciliated cells as well as the expansion of tuft and goblet cells. These compositional changes occur in an epithelium marked by increased prostaglandin activity, which likely further contributes to altered tissue function. IL-13-driven epithelial remodeling and PGE2-induced tissue effects may coordinately contribute to sinus opacification observed radiographically. With the increased use of type 2 directed biologics in clinical practice, future studies are needed to determine how such therapies alter the epithelial landscape.

## Data Availability

All data produced in the present study are available upon reasonable request to the authors

## Author contributions

All authors substantially contributed to the acquisition, analysis or interpretation of data for the manuscript and drafting, revising and critically reviewing the manuscript for important intellectual content. MEK, NNP, EKC, MAS, and EDG conceptualized the experiments. JGG, SDP, ANG recruited subjects and obtained epithelial brushes. MEK, NNP, CMM, and EDG performed experiments and analyzed data. MEK, CMM, and EDG synthesized the data and wrote the manuscript. All authors approved the final version of this manuscript.

## Acknowledgements

We gratefully acknowledge the support and participation of our patients.

## Figure Legends

*Supplementary Table 1: Unadjusted and adjusted statistics for correlations and box plots*

*Supplementary Table 2: Gene signature lists used for scoring*

